# Respiratory-Postural Responses to Singing: The Effects of Vocalisation and Tempo

**DOI:** 10.64898/2026.10.12.26364943

**Authors:** Nathan Healey, Elmar Kal, Alec Hewstone, Rayner Seavey, Grant Peterson, Keir E.J Philip, Ben Ainsworth, Adam Lewis

## Abstract

**Purpose:** Impaired balance and fall risk in individuals with respiratory conditions highlight the importance of respiratory-postural integration during upright activity. However, breathing and postural control are often studied separately, limiting understanding of their interaction. Singing imposes concurrent ventilatory and trunk control demands, yet respiratory-postural responses during standing singing remain unclear. We examined the effects of vocalisation and tempo on respiratory-postural responses during standing, and whether age and singing experience moderated these responses.

**Methods:** Eighty healthy younger (18-35 yr; n=35) and older (≥60 yr; n= 45) adults completed rhythmically paced speaking and singing tasks at slow, normal, and fast tempos while standing on a force platform. Respiratory rate (RR), mediolateral and anteroposterior sway, peripheral oxygen saturation, and perceived breathlessness were measured. Generalised estimating equations assessed vocalisation, tempo, and their interaction, with moderation by age and singing experience.

**Results:** RR showed a vocalisation x tempo interaction (p < .001), with higher RR during slow speaking and greatest increases during fast singing. Singing increased postural sway relative to speaking, particularly mediolaterally (p < .001), with tempo-related increases in anteroposterior sway (p < .001). Older adults exhibited greater postural sway overall (p values ≤ .040) but similar task-dependent modulation to younger adults. Singing experience attenuated RR increases during fast singing (p = .033) and was associated with greater mediolateral sway (p = .004). Perceived breathlessness increased with tempo but was not associated with vocalisation; SpO_2 w_as unchanged.

**Conclusions:** Standing singing was associated with tempo-dependent respiratory and postural responses. Age and singing experience showed distinct associations with these responses. Task-dependent respiratory-postural coordination appeared preserved among participants.

## 1. Introduction

Balance and breathing are both essential for everyday upright function, yet they are often studied as separate systems (Hodges et al. 2002; Janssens et al. 2024). This separation is clinically relevant because groups with chronic respiratory conditions often have impaired balance and an increased risk of falls (Loughran et al. 2020; Núñez-Cortés et al. 2022). In chronic obstructive pulmonary disease (COPD), for example, balance is poorer than in healthy peers, and falls are common (Loughran et al. 2020; Oliveira et al. 2021). Such impairments are likely multifactorial, with general deconditioning, reduced muscle strength, lower physical activity, and impaired coordination all contributing over time (Neder et al. 2019; O’Donnell et al. 2017). However, emerging evidence suggests that respiratory and postural systems may also interact more directly during upright activity (Hodges et al. 2002; Janssens et al. 2024). This has important clinical relevance, as respiratory rehabilitation now aims to improve not only symptoms and exercise tolerance, but also functional performance in everyday balance tasks (Rodrigues et al. 2020; Troosters et al. 2023).

Breathing and postural control are closely linked. During standing, breathing creates internal perturbations that must be managed by continuous postural adjustments (Hodges et al. 2002; Ivanenko and Gurfinkel 2018). The diaphragm, abdominal muscles, rib cage muscles, and other trunk stabilisers support both ventilation and trunk control (Hodges and Gandevia 2000; Kolar et al. 2012). Experimental evidence suggests that respiratory demand, trunk muscle coordination, and airway/glottal control can all influence postural control during upright activity (Hodges et al. 2002; Massery et al. 2013). A recent systematic review further showed that acute increases in respiratory demand can alter centre-of-pressure motion during standing, especially when respiratory loading is substantial (Janssens et al. 2024). Even so, the evidence base remains limited. Breathing and posture have still largely been studied as separate systems rather than as interacting components of functional upright behaviour (Hodges et al. 2002; Janssens et al. 2024).

This gap is particularly relevant because singing-based interventions are increasingly used to support breathing control in people with chronic respiratory disease (Lewis et al. 2021). Singing for Lung Health and related approaches have been associated with improvements in dyspnoea, breathing control, exercise capacity, and aspects of health-related quality of life, but the mechanisms underlying these benefits remain incompletely understood (Kaasgaard et al. 2022; Lewis et al. 2016; Lewis et al. 2021; Philip et al. 2024; Philip et al. 2020). Singing shares key features with breathing-focused physiotherapy and respiratory muscle training, including controlled inspiration, prolonged expiration or phonation, airflow regulation, and coordinated thoracoabdominal movement (Lewis et al. 2021; McConnell 2013). Therefore, singing may offer both a clinically relevant intervention and a useful model for studying respiratory control during upright tasks (Kaasgaard et al. 2022; Lewis et al. 2021).

Compared with conversational speech, singing places distinct demands on the respiratory system (Salomoni et al. 2016). Singing involves prolonged phonation, altered inspiratory timing, and greater regulation of expiratory airflow than speaking (Fuchs and Rochet-Capellan 2021; Hixon 1987; Philip et al. 2021). The respiratory demands of singing are also likely to depend on the temporal structure of the task, as breathing behaviour is adapted to rhythmic constraints (Salomoni et al. 2016). Studies in trained singers suggest that these demands are supported by task-specific breathing strategies, including modulation of expiratory flow, lung volume management, and coordinated rib cage-abdominal behaviour (Salomoni et al. 2016; Traser et al. 2022). However, most of this literature has focused on expert singers or seated and highly controlled laboratory tasks, limiting generalisability to upright functional activities and to individuals without formal singing experience (Lewis et al. 2021; Philip et al. 2021). As a result, it remains unclear how these respiratory strategies translate to upright task performance, where breathing occurs alongside postural control demands.

Furthermore, little is known about how respiratory and postural responses interact during upright vocal tasks such as singing. First, respiratory and postural responses to singing have rarely been examined concurrently, despite the inherently coupled nature of breathing, vocalisation, and upright stance (Attali et al. 2019; Hodges and Gandevia 2000). Additionally, changes in postural sway during vocal tasks do not necessarily reflect impaired balance. Increased movement variability may instead represent adaptive motor behaviour depending on task demands (Stergiou et al. 2006; Woollacott and Shumway-Cook 2002). Such movement may also reflect task-related behaviours and cultural norms associated with singing, including deliberate sway or movement with the music. Finally, it remains unclear whether these responses differ according to age or prior singing experience, both of which may influence task coordination and adaptation to increasing demands (Maki 1997; Prado et al. 2007; Ward et al. 2022).

Therefore, we hypothesised that (i) respiratory and postural responses would be modulated by the interaction between vocalisation and tempo, such that singing, particularly at faster tempos, would impose greater respiratory demand and be associated with increased postural sway relative to speaking, reflecting enhanced respiratory-postural coupling under temporal constraint; (ii) age would amplify these responses, with older adults demonstrating greater increases in postural sway and reduced capacity to adapt respiratory-postural coordination to increasing task demands; and (iii) Prior singing experience would moderate task responses, such that individuals with singing experience would show lower respiratory rate responses and attenuated changes in postural sway during demanding vocal tasks.

## 2. Methods

This laboratory-based study used a repeated-measures design to examine the effects of vocalisation and tempo on postural and physiological responses. Age group (younger vs older adults) and singing experience (singer vs non-singer) were between-participant factors, while vocalisation (speaking vs singing) and tempo (slow, normal, fast) were manipulated within participants. The study protocol was approved by Brunel University of London’s College of Health, Medicine and Life Science’s Research Ethics committee (35423-MHR-Feb/2022-38168-2) and pre-registered on ClinicalTrials.gov (NCT05350436).^1^

### 2.1 Sample

Community-dwelling adults were recruited from the general population. Younger adults were defined as aged 18-35 years and older adults as aged ≥60 years, consistent with World Health Organization criteria (Organisation 2025). Exclusion of intermediate ages (36-59) was intentional to maximise between-group contrast. All participants were required to stand independently for at least one minute and safely complete speaking and singing tasks while standing.

Exclusion criteria included diagnosed respiratory, neurological, or cardiac conditions requiring regular medication, mobility impairments affecting balance, pregnancy, speech or voice impairments limiting vocal task participation, a positive Coronavirus Disease 2019 (COVID-19) test within the previous 15 days, or a diagnosis of post-COVID-19 condition (long COVID). Only healthy volunteers were included to minimise confounding from comorbidities.

### 2.2 Group Allocation

Participants were allocated to one of four groups: younger adult singers, younger adult non-singers, older adult singers, and older adult non-singers. Singing experience was self-reported and defined as at least 5 years of regular participation in organised singing activities or training (e.g. choir, musical theatre group). Regular participation was defined as approximately weekly involvement.

### 2.3 Sample Size Considerations

Sample size was informed by prior experimental studies examining physiological and postural responses to singing and related tasks, which have typically employed samples in the range of approximately 20-40 participants in controlled repeated-measures designs (Peultier-Celli et al. 2022; Philip et al. 2021). Using G*Power (v3.1) for a repeated-measures within-between interaction (F-test), assuming a moderate effect size (f = 0.25), α = .05, power (1-β) = .80, correlation among repeated measures = 0.5, and ε = 1, the estimated required sample size is approximately N = 78.

### 2.4 Experimental Procedure

Each participant attended a single laboratory session lasting approximately 40 minutes. All testing was conducted in a controlled laboratory environment by trained researchers using standardised procedures. Participants were instructed to wear comfortable clothing and to avoid strenuous physical activity prior to testing.

On arrival, participants received a verbal explanation of the study procedures and provided written informed consent. Demographic information was collected, after which participants were familiarised with the force platform and the vocal tasks. The experimental session consisted of a series of non-vocal baseline balance conditions followed by vocalised balance conditions, all performed on a force plate (see below). Participants were instructed to maintain a comfortable upright stance and remain in position throughout each trial, without taking steps or altering their base of support. Vocalisation type and tempo were manipulated within participants, and condition order was randomised across participants to minimise order and fatigue effects (see Fig. 1 for participant flow diagram of procedure).

**Fig. 1.**
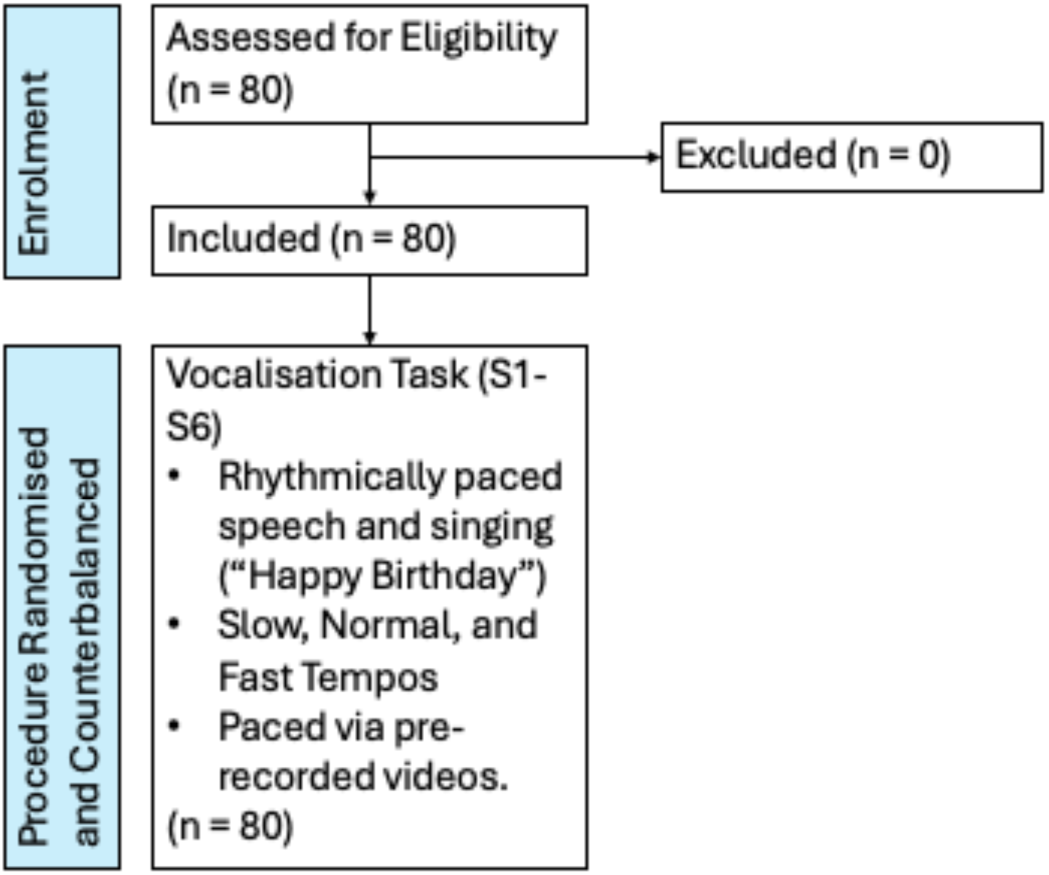
Flow Diagram Illustrating Participant Progression Through the Experimental Procedure.

### 2.5 Experimental Conditions

The experimental conditions are shown in Table 1. Primary analyses focused on the six rhythmically paced speaking or singing conditions (S1-S6; see Singing Balance Exercise Video.mp4), with quiet standing (B1; without vocalisation) used as the baseline comparison. All conditions were 30 seconds in duration.

**Table 1.**
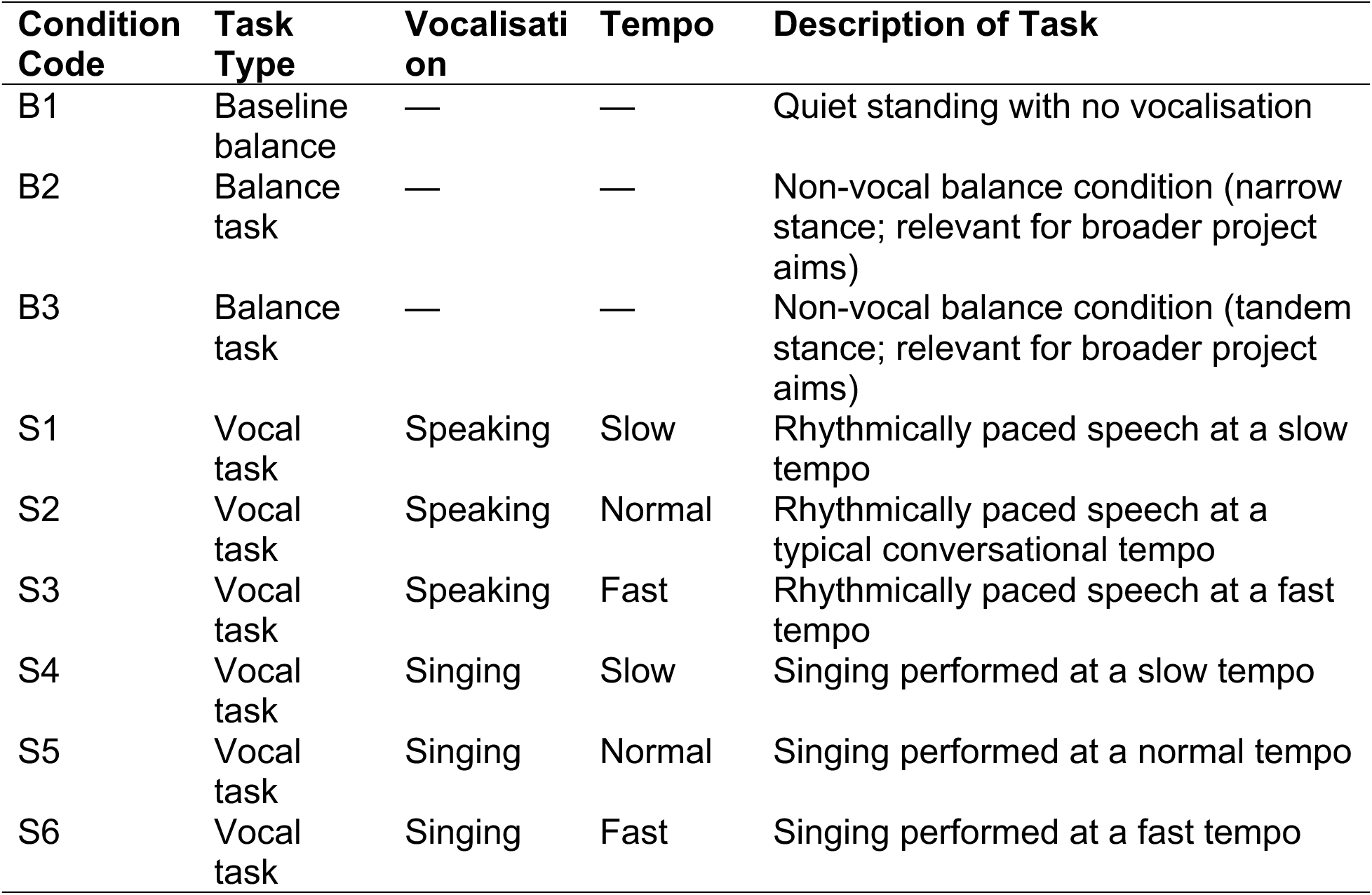
Experimental Conditions (all: 30 seconds)

| Condition Code | Task Type | Vocalisation | Tempo | Description of Task |
| --- | --- | --- | --- | --- |
| B1 | Baseline balance | — | — | Quiet standing with no vocalisation |
| B2 | Balance task | — | — | Non-vocal balance condition (narrow stance; relevant for broader project aims) |
| B3 | Balance task | — | — | Non-vocal balance condition (tandem stance; relevant for broader project aims) |
| S1 | Vocal task | Speaking | Slow | Rhythmically paced speech at a slow tempo |
| S2 | Vocal task | Speaking | Normal | Rhythmically paced speech at a typical conversational tempo |
| S3 | Vocal task | Speaking | Fast | Rhythmically paced speech at a fast tempo |
| S4 | Vocal task | Singing | Slow | Singing performed at a slow tempo |
| S5 | Vocal task | Singing | Normal | Singing performed at a normal tempo |
| S6 | Vocal task | Singing | Fast | Singing performed at a fast tempo |

### 2.6 Standing Balance Task

Participants stood barefoot on an AMTI Optima AccuSway force platform (500 x 500 mm; AMTI Inc., Watertown, MA, USA), looking straight ahead, with their arms by their sides and feet positioned at hip width (with positioning marked using tape to ensure consistency across trials).

### 2.7 Vocal Task

Prior to vocal tasks, participants completed a brief guided warm-up involving jaw mobility and breathing exercises to promote vocal comfort and consistency. Vocal tasks involved either rhythmically paced speech or singing of the Happy Birthday lyrics while standing on the force platform, ensuring that the speaking-versus-singing comparison used matched linguistic content. Tempos were externally paced via a pre-recorded audio track of a standardised model performance, identical across participants. Participants were instructed to synchronise their speech or singing with the recording throughout each condition.

### 2.8 Outcome Measures

The force platform sampled ground reaction forces and moments at 100 Hz. These were low-pass filtered offline with a bidirectional, second order Butterworth filter (cut-off frequency: 5 Hz), before estimating the centre-of-pressure coordinates using Matlab (MathWorks 2024). Postural sway was quantified as root-mean-square displacement (mm) in the anteroposterior (RMS-Y) and mediolateral (RMS-X) directions, for each condition.

Peripheral oxygen saturation (SpO_2_) was measured continuously using finger-based pulse oximetry, with the probe attached to the index finger of the non-dominant hand. Participants were instructed to keep the measured hand relaxed and still. Values were recorded at the end of each task.

Respiratory rate (RR) was assessed via direct observation to allow unobtrusive measurement during standing vocal tasks, where instrumentation may interfere with posture, phonation, or natural breathing behaviour. While this method lacks breath-by-breath resolution, it is well suited to capturing task-related breathing frequency under functional conditions. Importantly, respiratory rate was interpreted as a relative within-task indicator rather than an absolute measure of ventilatory output.

Perceived breathlessness was assessed immediately after each condition using the Modified Borg Dyspnoea scale (Borg 1998). Participants were familiarised with the scale prior to testing and received standardised instructions.

### 2.9 Statistical Analysis

Participants were classified into one of four groups, based on age and self-reported singing experience (as defined earlier): younger adult singers, younger adult non-singers, older adult singers, and older adult non-singers.

Analyses were performed using Generalised Estimating Equations (GEE) in SPSS (v30) to account for repeated measures. Participant identifier was specified as the clustering variable, and we used an exchangeable correlation structure and robust standard errors.

Primary analyses examined the effects of vocalisation (speaking vs. singing) and tempo (slow, normal, fast) as within-participant factors. Models included the main effects of vocalisation and tempo and their interaction (vocalisation x tempo). Separate models were fitted for each outcome variable (RR, RMS-X, RMS-Y, SpO₂, and Borg dyspnoea).

Secondary analyses examined whether these effects were moderated by participant characteristics. Two additional sets of models were fitted, incorporating either age group (younger vs. older adults) or singing experience (singers vs. non-singers) as a between-participant factor. These models retained the primary vocalisation x tempo structure and included the moderator as well as all relevant interaction terms (vocalisation x tempo, vocalisation x moderator, tempo x moderator, and vocalisation x tempo x moderator) to assess potential moderation of the primary effects.

Statistical significance was set at α = 0.05 using Wald χ² tests. Estimated marginal means with standard errors were reported. Holm-Bonferroni correction was applied to post-hoc comparisons.

## 3. Results

Full participant demographic characteristics are presented in Table 2. The dataset comprised of 80 participants and 480 trial observations distributed across six experimental conditions (S1-S6). Participants had a mean age of 51.4 ± 23.1 years (range = 21-80), and 63.7% were female. Mean participant height and body mass were 166.87 ± 8.9 cm and 70.3 ± 15 kg, respectively. Participants with prior singing experience comprised 38.8% of the sample and reported a mean singing history of 11.5 ± 18.2 years.

**Table 2.**
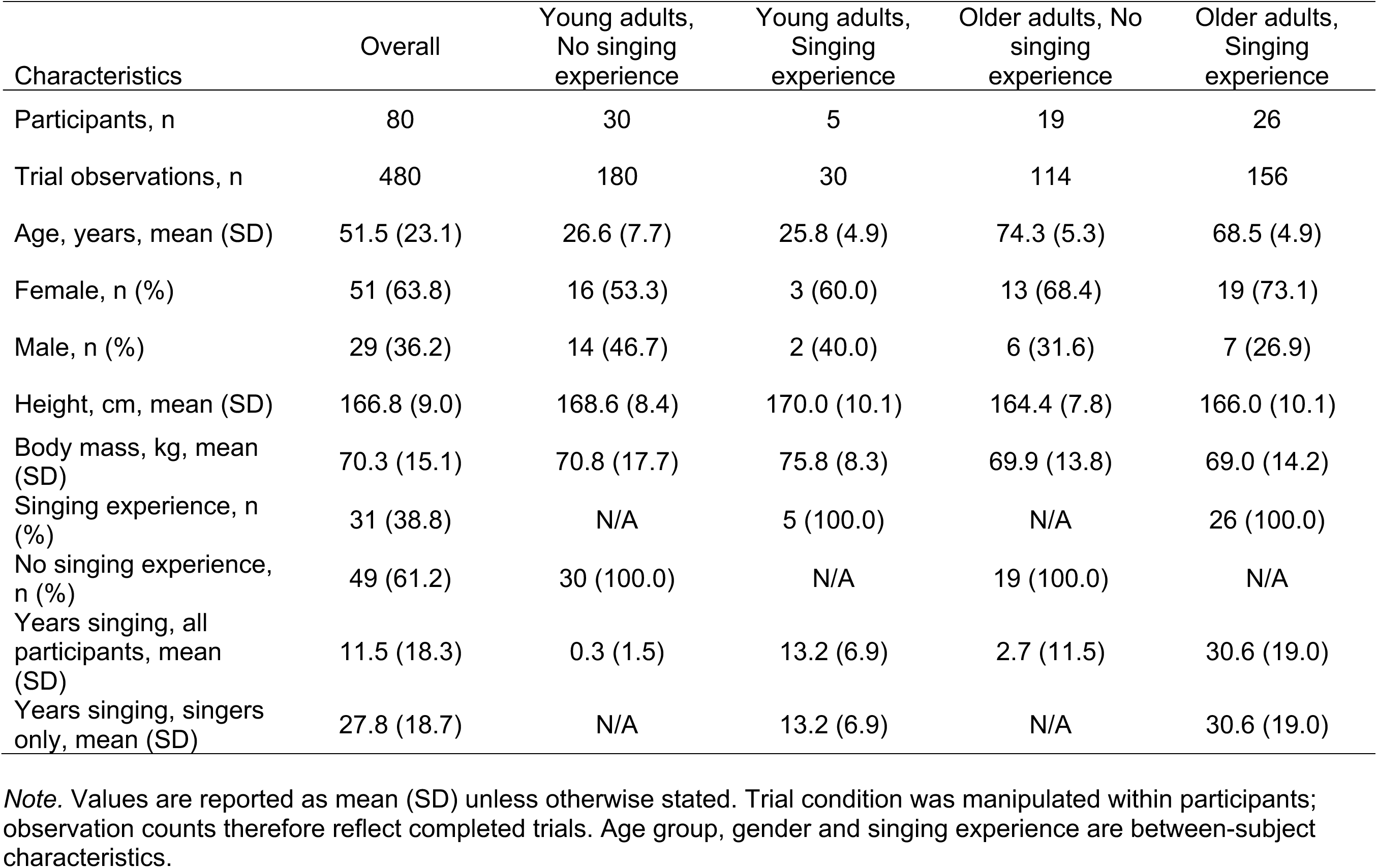
Demographic Characteristics and Trial Participation.

| Characteristics | Overall | Young adults,<br>No singing<br>experience | Young adults,<br>Singing<br>experience | Older adults, No<br>singing<br>experience | Older adults,<br>Singing<br>experience |
| --- | --- | --- | --- | --- | --- |
| Participants, n | 80 | 30 | 5 | 19 | 26 |
| Trial observations, n | 480 | 180 | 30 | 114 | 156 |
| Age, years, mean (SD) | 51.5 (23.1) | 26.6 (7.7) | 25.8 (4.9) | 74.3 (5.3) | 68.5 (4.9) |
| Female, n (%) | 51 (63.8) | 16 (53.3) | 3 (60.0) | 13 (68.4) | 19 (73.1) |
| Male, n (%) | 29 (36.2) | 14 (46.7) | 2 (40.0) | 6 (31.6) | 7 (26.9) |
| Height, cm, mean (SD) | 166.8 (9.0) | 168.6 (8.4) | 170.0 (10.1) | 164.4 (7.8) | 166.0 (10.1) |
| Body mass, kg, mean (SD) | 70.3 (15.1) | 70.8 (17.7) | 75.8 (8.3) | 69.9 (13.8) | 69.0 (14.2) |
| Singing experience, n (%) | 31 (38.8) | N/A | 5 (100.0) | N/A | 26 (100.0) |
| No singing experience, n (%) | 49 (61.2) | 30 (100.0) | N/A | 19 (100.0) | N/A |
| Years singing, all participants, mean (SD) | 11.5 (18.3) | 0.3 (1.5) | 13.2 (6.9) | 2.7 (11.5) | 30.6 (19.0) |
| Years singing, singers only, mean (SD) | 27.8 (18.7) | N/A | 13.2 (6.9) | N/A | 30.6 (19.0) |

*Note.* Values are reported as mean (SD) unless otherwise stated. Trial condition was manipulated within participants; observation counts therefore reflect completed trials. Age group, gender and singing experience are between-subject characteristics.

### 3.1 Primary Analysis: The Effects of Vocalisation and Tempo on Respiratory and Postural Outcomes

The overall model effects for respiratory, postural, and secondary outcomes are presented in Table 3 and 4, including the primary effects of vocalisation and tempo. As well as moderation analyses incorporating age group and singing experience. Estimated marginal means for these models are reported in Table 5.

**Table 3.**
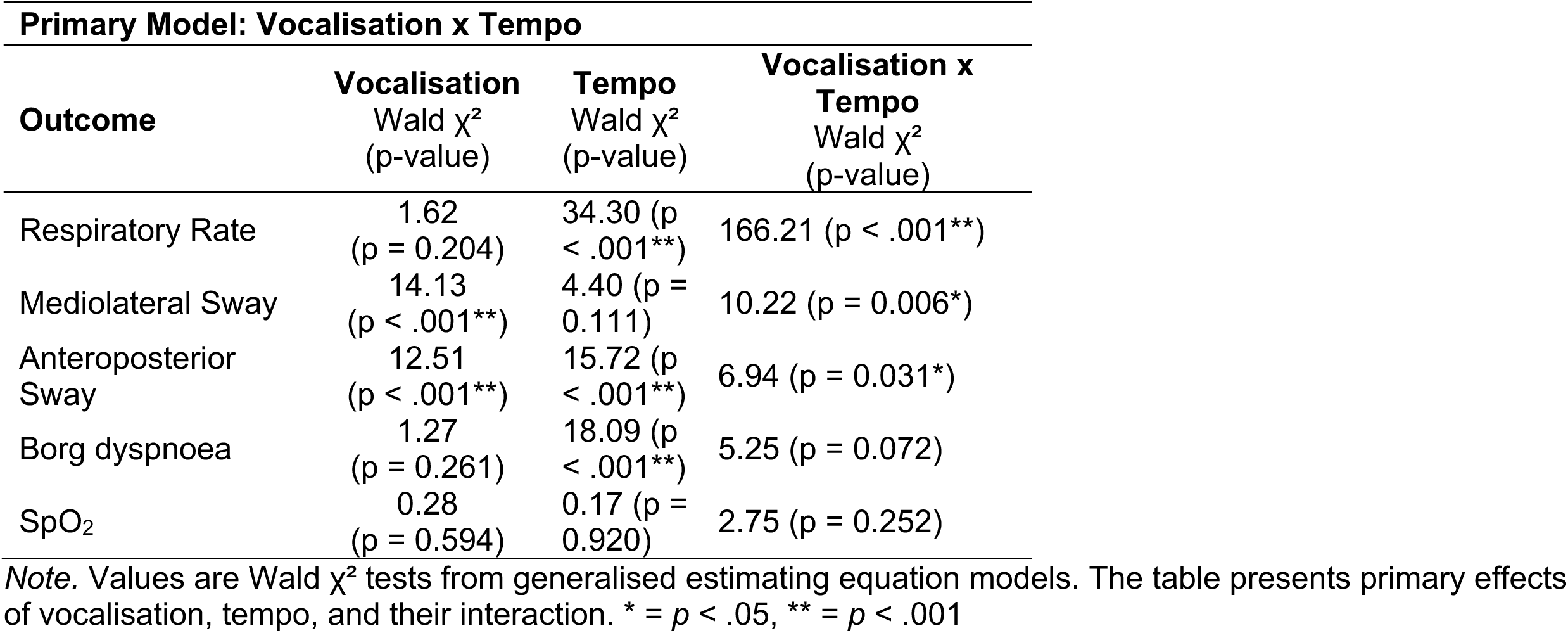
Summary of Generalised Estimating Equation for the Primary Model.

| **Primary Model: Vocalisation x Tempo** |  |  |  |
| --- | --- | --- | --- |
| **Outcome** | **Vocalisation**<br>Wald $\chi^2$<br>(p-value) | **Tempo**<br>Wald $\chi^2$<br>(p-value) | **Vocalisation x Tempo**<br>Wald $\chi^2$<br>(p-value) |
| Respiratory Rate | 1.62<br>(p = 0.204) | 34.30 (p < .001**) | 166.21 (p < .001**) |
| Mediolateral Sway | 14.13<br>(p < .001**) | 4.40 (p = 0.111) | 10.22 (p = 0.006*) |
| Anteroposterior Sway | 12.51<br>(p < .001**) | 15.72 (p < .001**) | 6.94 (p = 0.031*) |
| Borg dyspnoea | 1.27<br>(p = 0.261) | 18.09 (p < .001**) | 5.25 (p = 0.072) |
| SpO<sub>2</sub> | 0.28<br>(p = 0.594) | 0.17 (p = 0.920) | 2.75 (p = 0.252) |

*Note.* Values are Wald $\chi^2$ tests from generalised estimating equation models. The table presents primary effects of vocalisation, tempo, and their interaction. \* = $p < .05$ , \*\* = $p < .001$

**Table 4.**
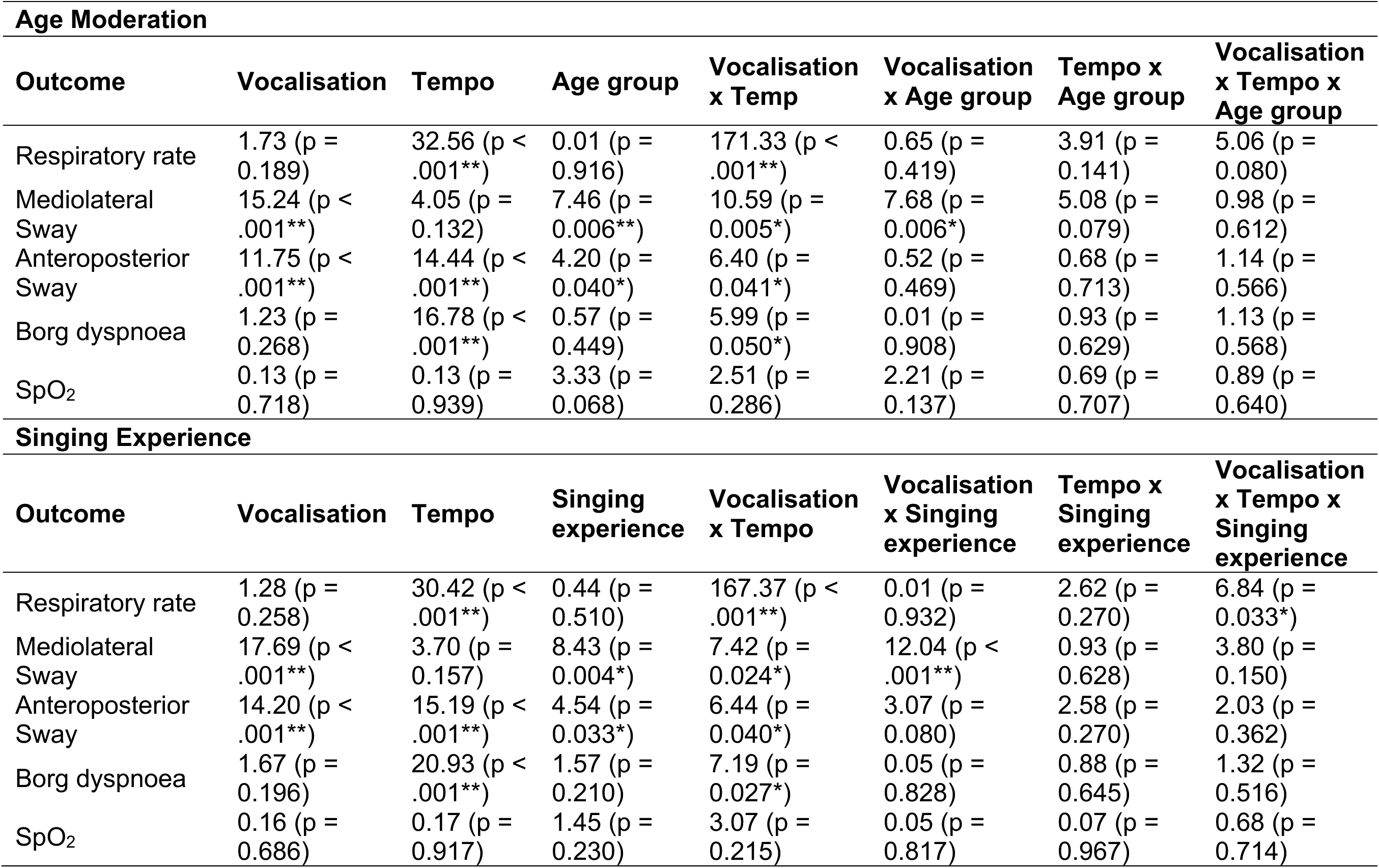
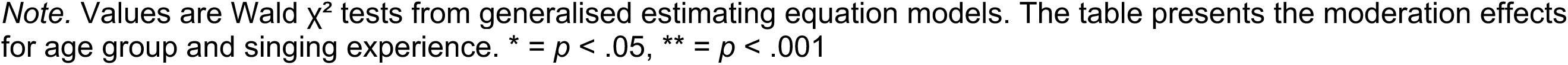
Summary of Generalised Estimating Equations for the Moderation of Age and Singing Experience.

**Table 5.**
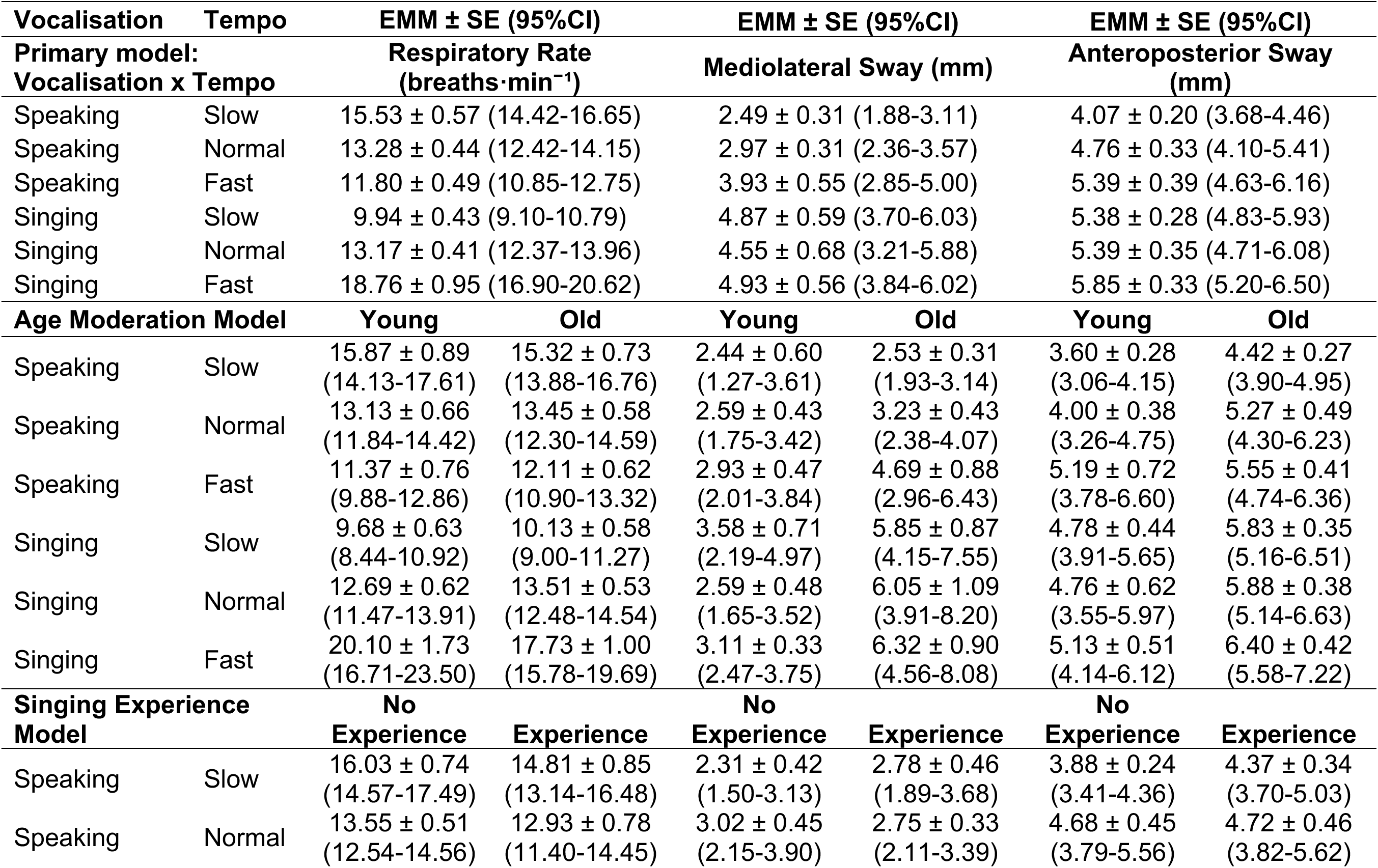
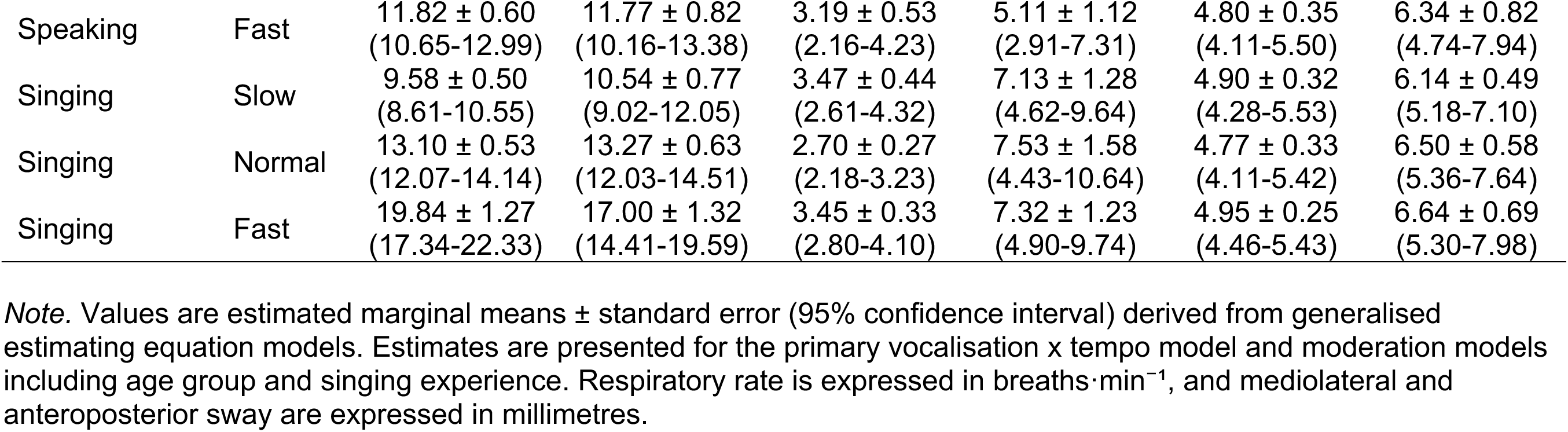
Estimated Marginal Means for Respiratory Rate, Mediolateral and Posteroanterior Sway Across Vocalisation and Tempo in the Primary, Age Moderation, and Singing-Experience Moderation Models.

#### 3.1.1 Respiratory Responses

RR showed a significant vocalisation x tempo interaction (p < .001), with a main effect of tempo (p < .001) but no main effect of vocalisation. Estimated marginal means indicated a crossover pattern. RR was higher during speaking at slow tempo, whereas rates were highest during singing at fast tempo. At normal tempo, RR was similar between conditions (Table 5; Fig. 2a).

**Fig. 2.**
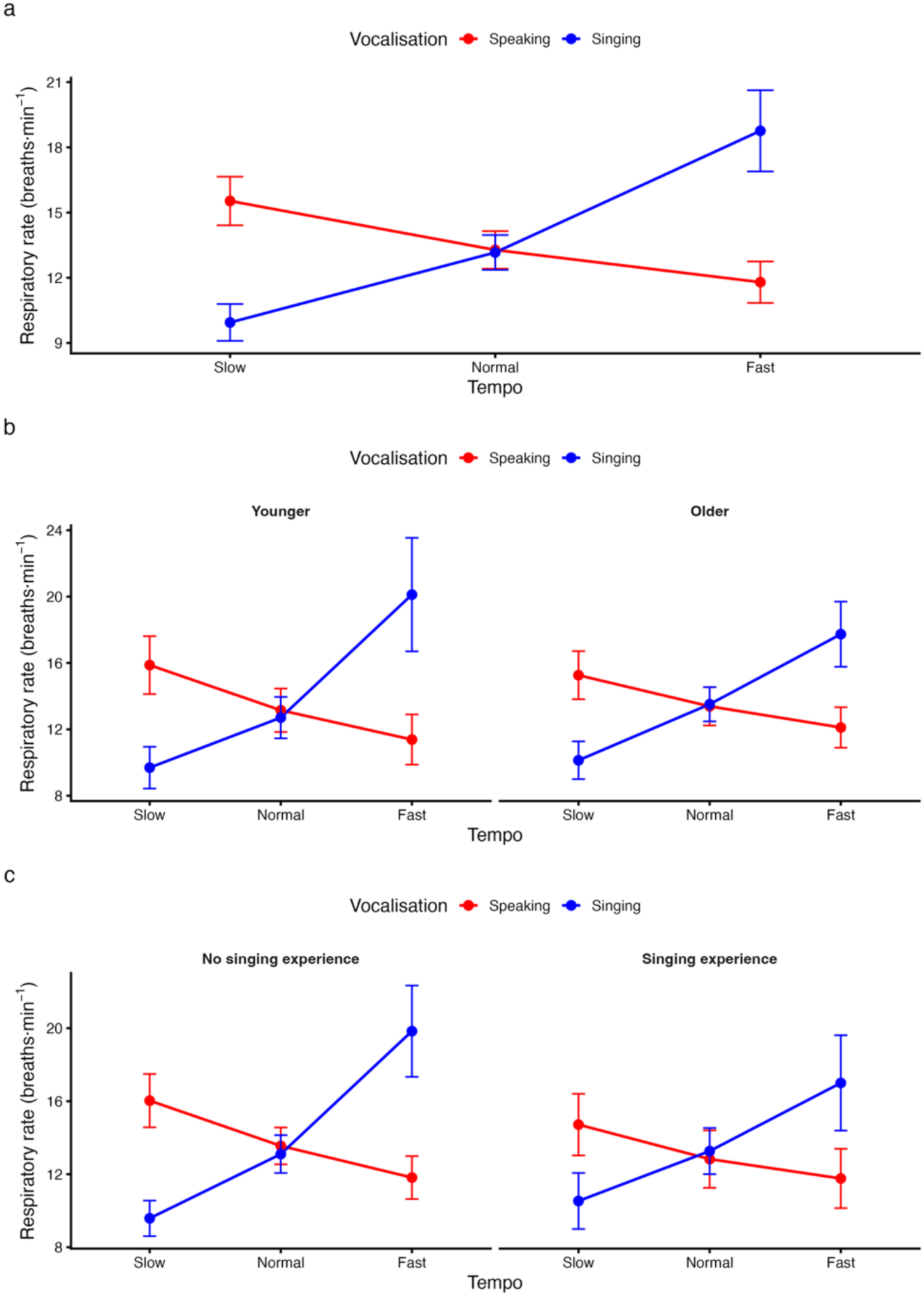
Difference in Type of Vocalisation and Tempo on Respiratory Rate. *Note.* (a) Primary vocalisation x tempo interaction for respiratory rate. (b) Effects stratified by age group. (c) Effects stratified by singing experience. Values represent estimated marginal means derived from generalised estimating equation models, with error bars indicating 95% confidence intervals. Respiratory rate is expressed in breaths·min^-1^.

#### 3.1.2 Postural Responses

For mediolateral sway amplitude, there was a significant main effect of vocalisation and a vocalisation x tempo interaction (both p = .006), with no main effect of tempo.

Mediolateral sway was consistently greater during singing than speaking across all tempos, with the largest difference observed at slow tempo. Collapsed across tempo, singing was associated with increased mediolateral sway by 1.65 mm (95% CI 0.79 to 2.51; Table 5; Fig. 3a).

**Fig. 3.**
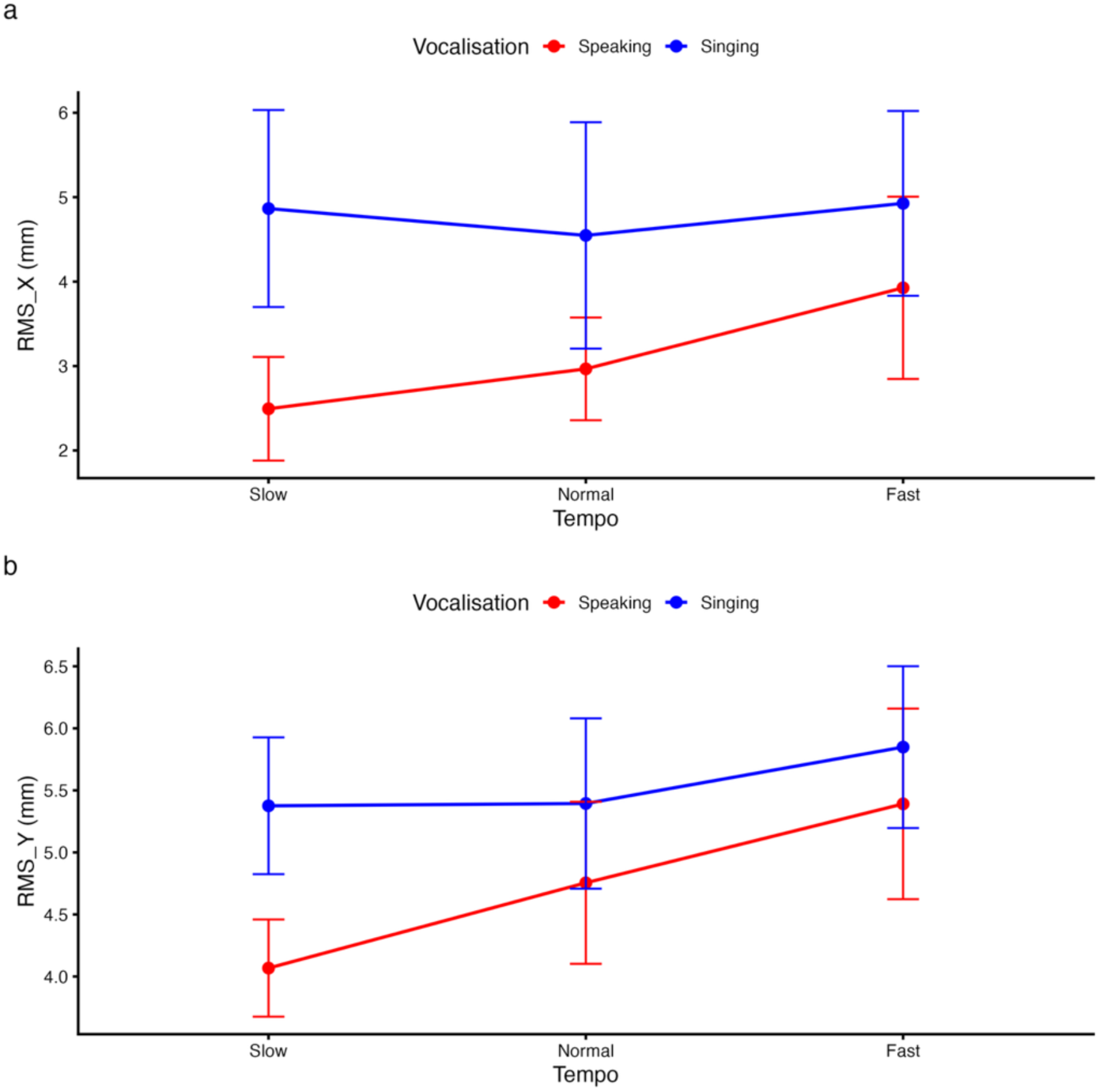
Differences in Type of Vocalisation and Tempo on Postural Sway. *Note.* (a) Mediolateral sway (RMS-X). (b) Anteroposterior sway (RMS-Y). Values represent estimated marginal means derived from generalised estimating equation models, with error bars indicating 95% confidence intervals.

For anteroposterior sway, a significant main effect of vocalisation and tempo was observed, alongside a vocalisation x tempo interaction (p = .031). Sway was greater at faster tempo and during singing across conditions. On average, singing was associated with 0.80 mm greater anteroposterior sway than speaking (95% CI 0.36 to 1.25), while fast tempo was associated with approximately 0.90 mm greater sway relative to slow tempo (Table 5; Fig. 3b).

### 3.2 Responses to Vocalisation and Tempo in Younger and Older Adults

#### 3.2.1 Respiratory responses

Including age group did not alter the primary pattern of results. The vocalisation x tempo interaction remained significant (p < .001), with no main effect of age and no higher-order interactions. The crossover pattern was evident in both younger and older adults, although respiratory rate during fast singing was numerically greater in younger participants (Fig. 2b). No effects involving age were observed for Borg dyspnoea or SpO_2._

#### 3.2.2 Postural responses

For mediolateral sway, significant effects of vocalisation and age were observed, along with vocalisation x tempo and vocalisation x age interactions (p = .006). Older adults exhibited greater sway overall (mean difference 1.91 mm, 95% CI 0.54 to 3.28), and the increase associated with singing was more pronounced in this group (Table 5; Figure 4a).

**Fig. 4.**
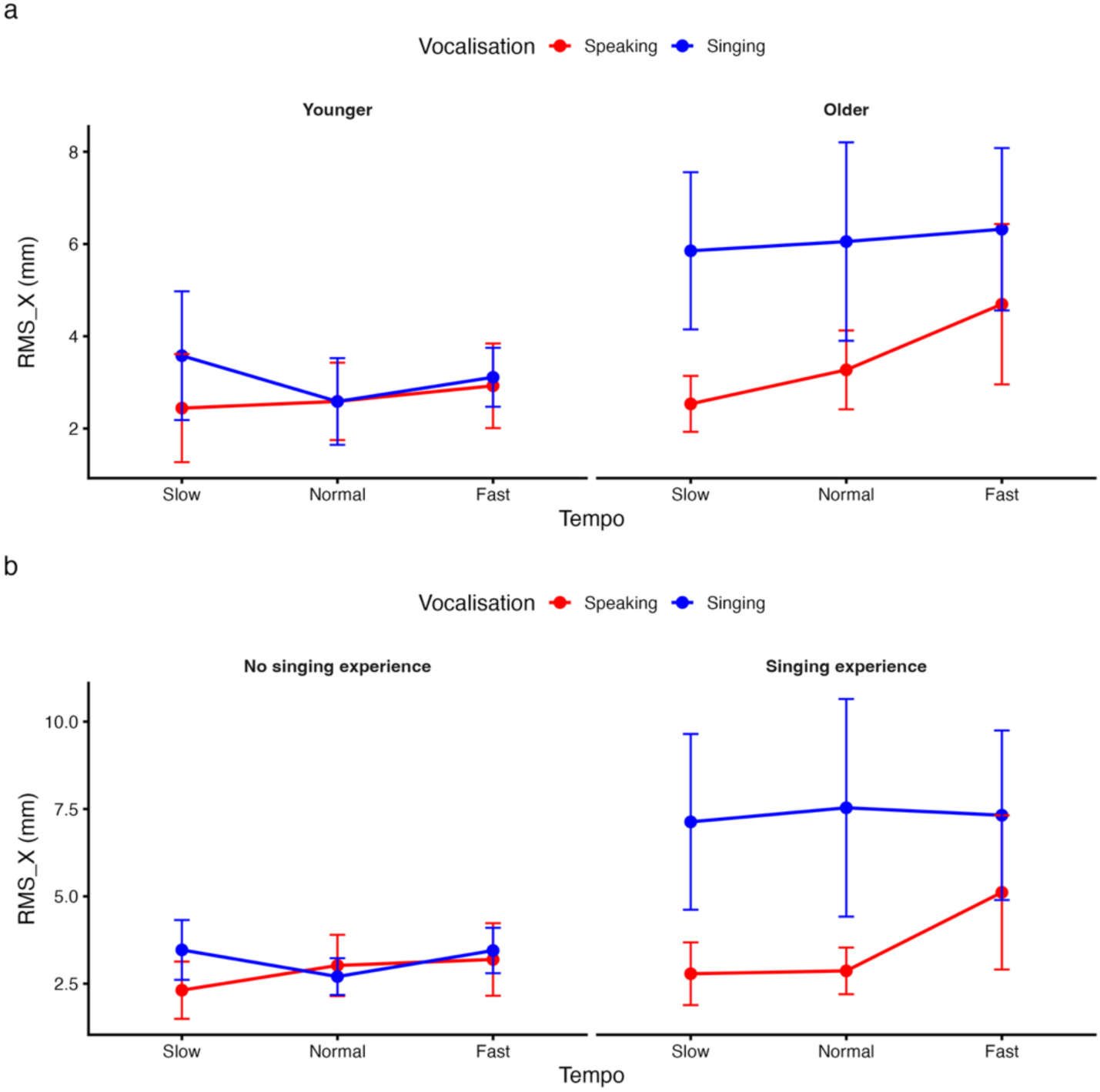
Moderating Effects of Age and Singing Experience on Mediolateral Sway (RMS-X) *Note.* (a) Effects stratified by age group. (b) Effects stratified by singing experience. Values represent estimated marginal means derived from generalised estimating equation models, with error bars indicating 95% confidence intervals.

For anteroposterior sway, vocalisation, tempo, and age showed significant main effects, but no interactions involving age were identified. Older adults demonstrated higher sway overall, while the anteroposterior sway during singing and at faster tempo was preserved.

### 3.3 Responses to Vocalisation and Tempo by Singers and Non-Singers

#### 3.3.1. Respiratory responses

A significant vocalisation x tempo x singing experience interaction was observed (p = .033). The crossover pattern remained consistent across groups. However, during fast singing, participants without singing experience exhibited higher RR than those with experience, whereas responses during speaking were similar between groups (Table 5; Fig. 2c). No significant associations involving singing experience were observed for Borg dyspnoea or SpO_2._

#### 3.3.2. Postural responses

For RMS-X, significant main effects of vocalisation and singing experience were observed, along with interactions between vocalisation and both tempo and singing experience (p = .004). Singing experience was associated with substantially greater sway during singing compared with non-singers, with between-group differences of approximately 4 mm (Table 5; Fig. 4b).

For RMS-Y, vocalisation, tempo, and singing experience showed significant main effects, with no interactions involving singing experience. Anteroposterior sway was greater during singing and faster tempos in both groups.

## 4. Discussion

This study examined respiratory and postural responses during upright speaking and singing tasks performed at different tempos, and whether these responses differed by age group or singing experience. The main findings were that RR varied according to the interaction between vocalisation type and tempo, with higher values during slow speaking and the highest values during fast singing. Postural sway was generally greater during singing than speaking, with mediolateral sway showing differences between vocalisation types and anteroposterior sway also varying by tempo. Age group was not associated with differences in respiratory rate responses, although older adults showed greater postural sway overall. Singing experience was associated with lower RR values during fast singing and greater mediolateral sway during singing. Perceived breathlessness varied by tempo but not vocalisation type, while SpO_2 r_emained stable across conditions. Together, these findings indicate that increasing task demand shifts coordination toward integrated respiratory-postural strategies, with efficiency achieved through increasing rather than constraining movement variability.

The interaction in respiratory rate likely reflects temporal constraints imposed by the vocal task. Singing requires coordination of inspiration with prolonged, rhythmically structured expiration, limiting inspiratory time as tempo increases (Philip et al. 2021; Salomoni et al. 2016). Accordingly, slower tempos permit longer expiratory phases and lower breathing frequencies, whereas faster tempos necessitate increased respiratory rate. These findings extend prior work by suggesting that respiratory frequency responses are shaped by the interaction between task type and temporal constraint, rather than solely reflecting metabolic demand (Salomoni et al. 2016). This likely reflects differences in motor control organisation, with singing relying more heavily on learned, feedforward coordination of respiratory and laryngeal systems, whereas speech permits greater flexibility and reactive adjustment.

Lower RR responses in singers may indicate different breath-control strategies under demand. Increased breathing frequency is associated with ventilatory inefficiency and dyspnoea (Caviedes 2021; O’Donnell et al. 2017). Trained singers rely on modulation of expiratory flow, lung volume, and coordinated rib cage-abdominal motion to sustain ventilation (Salomoni et al. 2016; Traser et al. 2022). While respiratory rate alone does not directly quantify ventilatory efficiency, the absence of increased breathlessness or oxygen desaturation alongside lower frequency values suggests that singers may have completed the task through alternative strategies, such as greater tidal volume or improved expiratory control, rather than reduced ventilatory demand.

The present findings therefore show that these strategies extend to upright functional tasks, consistent with reported benefits of singing-based interventions (Lewis et al. 2021; Yuen and Ma 2024). This suggests that the observed RR pattern may reflect active breath-control strategies rather than reduced ventilatory demand, although respiratory efficiency was not directly measured. Equally, RR during singing may also have been shaped by the phrasing and tempo of the song. Breathing opportunities occur at relatively fixed points and become closer together as tempo increase (Salomoni et al. 2016).

Postural responses may also be consistent with altered respiratory-postural coupling during singing. Breathing generates internal perturbations that must be accommodated by changes in postural control (Hodges et al. 2002), with mediolateral sway particularly sensitive to trunk-mediated adjustments (Mujdeci et al. 2016; Prado et al. 2007). Increased sway during singing, particularly at faster tempos, therefore, could reflect adaptive coordination rather than instability, consistent with motor control frameworks in which variability supports task performance (Stergiou et al. 2006; Woollacott and Shumway-Cook 2002). Further studies using controlled postural perturbations are needed to determine whether greater sway during singing translates into improved, reduced, or unchanged capacity to respond to small external balance disturbances. Age-related increases in sway align with established declines in postural control under dual-task conditions (Maki 1997; Ward et al. 2022), but preserved task-related modulation indicates that older adults retain the capacity to adjust their postural control in response to changing respiratory demands. This suggests that ageing influences the efficiency, rather than the organisation, of coordination in response to increased breathing control demands. Notably, singing experience was associated with greater mediolateral sway, challenging the view that improved performance is characterised by reduced movement. Instead, skilled behaviour may be associated with flexible, task-specific variability (Stergiou et al. 2006). Increased trunk motion in singers may facilitate fine control of airflow and subglottal pressure, supporting efficient phonation under temporal constraint. The combination of lower respiratory rate and greater postural movement may be consistent with coordinated respiratory-postural strategies rather than a simple stabilisation response.

### 4.1 Implications

These findings provide mechanistic insight into respiratory and postural behaviour during singing and support the conceptualisation of singing as a respiratory control task performed under postural load, rather than as a purely expressive activity (Lewis et al. 2021). The lower RR values observed in singers may be relevant to respiratory rehabilitation, where singing-based approaches often aim to support breath control and symptom management. However, RR should be interpreted cautiously, as higher breathing frequency is not necessarily maladaptive and may reflect an appropriate response to task demand rather than a contributor to dyspnoea or activity limitation. Singing-based interventions have been associated with improvements in breathing control, dyspnoea, and functional outcomes, particularly in older adults and individuals with chronic respiratory disease (Kaasgaard et al. 2022; Lewis et al. 2016; Lewis et al. 2021; Philip et al. 2024; Philip et al. 2022; Philip et al. 2026; Yuen and Ma 2024). The present findings may inform future research on singing-based interventions by showing that upright singing is associated with distinct respiratory and postural response patterns. However, clinical implications should be drawn cautiously. The sample comprised healthy adults, the task differed substantially from structured respiratory rehabilitation or Singing for Lung Health interventions, and RR differences were likely shaped by musical phrasing as well as physiological demand. Similarly, greater postural sway during singing may reflect behavioural responses to music, confidence, or task familiarity, rather than respiratory-postural control alone. These findings therefore emphasise the importance of interpreting balance outcomes during singing in relation to respiratory strategy, musical structure, and movement behaviour (Hodges et al. 2002; Prado et al. 2007; Stergiou et al. 2006).

### 4.2 Strengths and Limitations

A key strength of this study is the integrated assessment of respiratory and postural responses during functional vocal tasks performed in upright standing. Manipulation of tempo enabled controlled increases in task demand, and the inclusion of physiological and perceptual measures strengthened interpretation of respiratory efficiency. By focusing on experience-dependent differences in acute responses rather than solely on professional singers or intervention outcomes, the study extends current research and highlights respiratory rate as a pragmatic, task-compatible measure of breathing pattern during upright vocal performance.

The cross-sectional design limits causal inference, meaning that singing experience cannot be definitively identified as the source of the observed adaptations. It is also possible that individuals with inherently more efficient respiratory-postural coordination are more likely to engage in or maintain singing activities. The use of a healthy adult sample may limit generalisability to clinical populations with respiratory or balance impairment. Additional demographic information including ethnicity and socioeconomic status would also have been useful for assessing representativeness of the sample (Buttery et al. 2022; Kenny et al. 2026). While respiratory rate does not capture tidal volume or minute ventilation, it was selected as a functionally meaningful measure compatible with upright vocal tasks. Finally, centre-of-pressure measures reflect global postural behaviour but cannot resolve segmental or muscular coordination, warranting further investigation using kinematic or electromyographic approaches.

## 5. Conclusions

Standing singing imposes task-dependent respiratory and postural demands that are strongly influenced by tempo, age, and prior experience. Singing experience was associated with lower respiratory rate values under increasing temporal demand, while greater mediolateral postural motion during singing may reflect task-specific movement strategies rather than instability alone. These findings emphasise the importance of respiratory-postural integration in understanding the functional effects of singing and provide a mechanistic basis for future work examining training effects and clinical translation.

## Acknowledgements

We would like to thank all the participants who volunteered their time to participate in this study. Furthermore, we would like to thank all the MSc Physiotherapy dissertation students for their work on this group-based project.

## Declarations

## Author Contributions

Adam Lewis and Elmar Kal were responsible for setting up the project and supervising the MSc physiotherapy students, including Alec Hewstone and Raynor Seavey who contributed to the study design and completed the data collection for this project. Keir EJ Philip contributed to project design. Grant Peterson contributed to the development of the vocal task instruction video. Sally Goodworth provided consultation on the study methodology. Nathan Healey was responsible for analysis of the data and write up of the paper. All authors reviewed draft manuscripts.

## Ethical Approval

This study was performed in line with the principles of the Declaration of Helsinki. Ethical approval was granted by Brunel University of London’s College of Health, Medicine and Life Science’s Research Ethics committee (35423-MHR-Feb/2022-38168-2).

## Consent to Participate

Informed consent was obtained from all participants included in the study. All participants provided consent to publish.

## Data Availability

Anonymised data may be shared on reasonable request.

## Abbreviations

COPD: Chronic Obstructive Pulmonary Disease
COVID-19: Coronavirus Disease 2019
GEE: Generalised Estimating Equation
RMS-X: Mediolateral Sway
RMS-Y: Anteroposterior Sway
RR: Respiratory Rate
SpO_2_: Oxygen Saturation

## Footnotes

1 Please note that the trial registration included some task conditions (conditions B2, B3, see Table 1 and main text) and outcomes that are not reported on as part of this paper, as these were not directly relevant to our current aims and analyses.

